# Longitudinal multi-omics study reveals common etiology underlying association between plasma proteome and BMI trajectories in adolescent and young adult twins

**DOI:** 10.1101/2023.06.28.23291995

**Authors:** Gabin Drouard, Fiona A. Hagenbeek, Alyce Whipp, René Pool, Jouke Jan Hottenga, Rick Jansen, Nikki Hubers, Aleksei Afonin, BIOS Consortium, BBMRI-NL Metabolomics Consortium, Gonneke Willemsen, Eco J. C. de Geus, Samuli Ripatti, Matti Pirinen, Katja M. Kanninen, Dorret I. Boomsma, Jenny van Dongen, Jaakko Kaprio

## Abstract

**Background:** The influence of genetics and environment on the association of the plasma proteome with body mass index (BMI) and changes in BMI remain underexplored, and the links to other omics in these associations remain to be investigated. We characterized protein-BMI trajectory associations in adolescents and adults and how these connect to other omics layers.

**Methods:** Our study included two cohorts of longitudinally followed twins: FinnTwin12 (*N*=651) and the Netherlands Twin Register (NTR) (*N*=665). Follow-up comprised four BMI measurements over approximately 6 (NTR: 23-27 years old) to 10 years (FinnTwin12: 12-22 years old), with omics data collected at the last BMI measurement. BMI changes were calculated using latent growth curve models. Mixed-effects models were used to quantify the associations between the abundance of 439 plasma proteins with BMI at blood sampling and changes in BMI. The sources of genetic and environmental variation underlying the protein abundances were quantified using twin models, as were the associations of proteins with BMI and BMI changes. In NTR, we investigated the association of gene expression of genes encoding proteins identified in FinnTwin12 with BMI and changes in BMI. We linked identified proteins and their coding genes to plasma metabolites and polygenic risk scores (PRS) using mixed-effect models and correlation networks.

**Results:** We identified 66 and 14 proteins associated with BMI at blood sampling and changes in BMI, respectively. The average heritability of these proteins was 35%. Of the 66 BMI-protein associations, 43 and 12 showed genetic and environmental correlations, respectively, including 8 proteins showing both. Similarly, we observed 6 and 4 genetic and environmental correlations between changes in BMI and protein abundance, respectively. *S100A8* gene expression was associated with BMI at blood sampling, and the *PRG4* and *CFI* genes were associated with BMI changes. Proteins showed strong connections with many metabolites and PRSs, but we observed no multi-omics connections among gene expression and other omics layers.

**Conclusions:** Associations between the proteome and BMI trajectories are characterized by shared genetic, environmental, and metabolic etiologies. We observed few gene-protein pairs associated with BMI or changes in BMI at the proteome and transcriptome levels.

## Introduction

In recent decades, the prevalence of obesity has been increasing [1], and it is predicted that almost one-fourth of the world’s population will be affected by obesity in 2035 [2]. The co-morbidities related to obesity include a wide range of high-prevalence diseases, including type 2 diabetes and cardiovascular disease [3–5], making it a major public health concern.

Body mass index (BMI) is commonly used as a marker of obesity and body fat, the former being defined as BMI >30 kg.m^-2^. Both genetics and the environment influence BMI, with twin studies estimating ∼75% of its variance attributable to genetic factors in young adults [6]. Multiple proteins and peptides, such as the hormones leptin, ghrelin, and resistin, take part in the complex processes of regulating energy balance, disturbances of which can induce obesity or anorexia [7,8]. High-throughput technologies have revolutionized obesity research, and omics data are key to an in-depth functional understanding of obesity [5]. An omics technique successful in investigating obesity-related co-morbidities is proteomics, which comprises the large-scale study of proteins [9]. For example, beyond the risk of general obesity in the development of diabetes, plasma protein markers of abdominal fat distribution are associated with an increased risk of diabetes [10]. Mendelian randomization identified unidirectional and bidirectional (i.e., protein-to-BMI and/or BMI-to-protein) causal associations of plasma proteins with BMI [11,12], signifying a direct imprint of the proteome on obesity, and vice versa. Mendelian randomization leverages genetic variants that influence protein levels and BMI, but little is known about how much of the association between protein levels and BMI is due to genetic and environmental influences.

Multi-omics approaches that combine multiple omics layers in a single analysis better characterize the underlying biology of complex diseases than single-omics approaches [13,14]. The few multi-omics studies conducted for obesity demonstrated great potential in acquiring a better understanding of obesity, notably when coupling transcriptomic and metabolomic data [15,16]. Specifically, one study showed that some metabolite and gene expression associations with BMI were mediated by the epigenome [15], while another study observed associations of weight gain with changes in metabolite levels and blood cell function [16]. Among twin pairs discordant for BMI, weight differences were associated with lipidomics independent of genetic influences [17]. Overall, only a few multi-omics studies quantified biomolecules (i.e., metabolites or proteins) and omics stability in relation to participants’ post-intervention weight changes [18–20].

Weight development and obesity in adolescents and children from a multi-omics perspective is largely underexplored. The large inter- and intra-individual variability in the proteome, metabolome, and transcriptome in children [21] holds great promise for identifying obesity biomarkers across omics layers. Better identification of biomarkers for obesity in children and adolescents may allow both prevention of its onset in this age group and in adults, as childhood obesity increases the likelihood of adult obesity [22]. Longitudinal designs can help capture factors associated with weight change, and thus better predict individuals at risk for obesity. A few longitudinal single-omic studies have explored the associations between changes in BMI with proteomic [23–26] or metabolomic data [27]. While the studies involving proteomic data are mainly based on adult populations, one study to investigate the association between weight change and metabolites was conducted in children [28]. No one, to our knowledge, has conducted longitudinal multi-omics studies of weight change and BMI including proteomics data in children or adolescent populations yet.

Plasma proteins are associated with BMI in adults, where some plasma proteins were shown to be causally influencing increases in BMI, while other plasma proteins were causally influenced by higher BMI [11,12]. As less is known about these associations in adolescence and for changes in BMI, the primary objective of the current study was to identify plasma proteins associated with BMI at blood sampling and changes in BMI during adolescence and in adulthood (Figure 1). Here, we refer to BMI at blood sampling and changes in BMI together as BMI trajectories. As changes in BMI in adolescents reflect substantial changes in lean body mass [29], observing an overlap of proteins associated with BMI changes during adolescence with those reported in the literature in adults could indicate whether proteins associated with BMI changes are due to changes in body fat or lean mass. We also elucidated the genetic and environmental sources underlying both the protein abundances and their correlations with BMI and changes in BMI. How genetic differences give rise to differences in plasma protein abundances and in BMI and changes in BMI is influenced by a complex interplay of multiple omics layers. To characterize the associations of multiple omics layers with BMI and changes in BMI, we next investigated whether gene expression of relevant protein-coding genes were associated with BMI trajectories in an external cohort. Finally, we linked identified proteins and their coding genes to plasma metabolites and polygenic risk scores (PRS) for obesity and coronary artery disease (CAD) using mixed-effect models and correlation networks.

**Figure 1:**
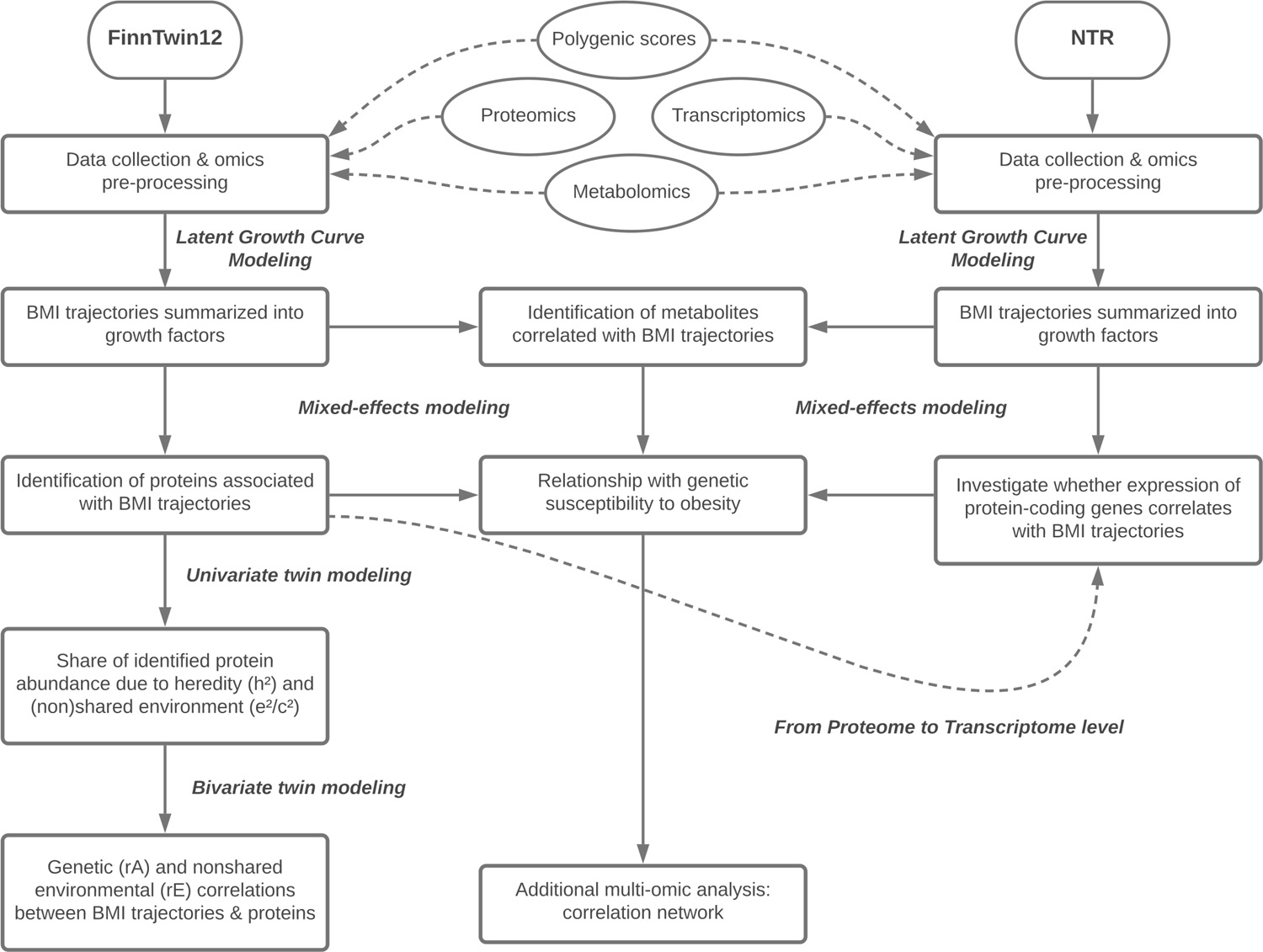
Study flowchart **legend:** The study was divided into two main sequential steps. First, growth factors were calculated and then association studies were performed. In the FinnTwin12 sample, univariate and bivariate twin modeling was performed to quantify the genetic and environmental sources underlying both the protein abundances and the correlations between proteins and BMI trajectories. Cross-omics associations were performed to bring a holistic perspective to the analyses. BMI: body mass index. NTR: Netherlands Twin Register.

## Methods

### Study population and longitudinal measurements

#### FinnTwin12

FinnTwin12 is a longitudinal cohort based on a population of Finnish twins born between 1983 and 1987 aimed at investigating adolescent behavioral development and health habits [30,31]. Participants were identified from the Central Finnish Population Register and completed four questionnaires (response rate range: 85–90%) at approximate ages 12, 14, 17, and 22. A subset of twins, referred to as intensive subset, was studied more intensively, with additional psychiatric interviews and questionnaires starting at the wave corresponding to age 14. Besides health, lifestyle, and psychological indicators, questionnaires collected up to four self-reported weight and height measures for these participants across the four waves, from which BMI values (kg.m^-2^) were calculated. At the 22-year assessment wave, 786 twins from the intensive subset participated in in-person assessments and provided venous blood plasma samples after overnight fasting. We assessed the validity of self-reports of weight and height at the 22-year assessment wave versus in-person measurements in a sample of 756 of these participants (see Additional file 2). Self-reported and measured values correlated strongly for weight (Pearson correlation: *r*=0.98) and height (*r*=0.99). From the blood samples, extensive omic data were derived, including proteomics, metabolomics, and genotyping to generate the PRS data used in the current study. Of these participants, 651 had complete longitudinal anthropometric measurements over the 10 years of follow-up and constituted the final FinnTwin12 sample (Table 1). The sample included 250 monozygotic twins (106 complete pairs) and 401 dizygotic twins (164 complete pairs). Height and weight across the waves can be found in the supplementary material (see Additional file 3, Table S1).

**Table 1:** Descriptive Statistics of the FinnTwin12 and NTR Cohorts **Legend:** Four longitudinal measurements with ∼10 and ∼6 years of follow-up were collected in the FinnTwin12 and NTR cohorts, respectively. Time points marked with (*) included blood sampling from which proteomic, metabolomic, and polygenic risk score data were derived. Intercepts and slopes were obtained by Latent Growth Curve Models, in a logarithmic scheme for FinnTwin12 and linear scheme for NTR. Final samples included 651 participants (female: 59%) for FinnTwin12 and 665 (female: 69%) for NTR. In FinnTwin12, BMI at baseline and last BMI measurement (wave 22) ranged 11.0-30.1 and 17.2-42.0 respectively. In NTR, BMI at baseline and last BMI measurement (bb1) ranged 15.2-40.3 and 15.8-51.3 respectively. BMI is expressed as kg/m2 and age as years. NTR: Netherlands Twin Register. N: Number of participants without missing value. sd: standard deviation.

| Timepoint | Cohort | N (% female) | Body mass index (BMI) |  |  |  | Age |  |
| --- | --- | --- | --- | --- | --- | --- | --- | --- |
|  |  |  | Male participants | sd | Female participants | sd |  |  |
| Wave 12 | FinnTwin12 | 651 (59%) | 17.7 | 2.8 | 17.6 | 2.4 | 11.4 | 0.3 |
| Wave 14 | FinnTwin12 | 651 (59%) | 19.5 | 3 | 19.5 | 2.6 | 14.0 | 0.1 |
| Wave 17 | FinnTwin12 | 651 (59%) | 21.8 | 2.9 | 21.2 | 2.8 | 17.6 | 0.2 |
| (*) Wave 22 | FinnTwin12 | 651 (59%) | 23.9 | 3.4 | 22.6 | 3.6 | 22.4 | 0.7 |
| Survey 5 | NTR | 412 (71%) | 22.0 | 2.9 | 21.2 | 2.7 | 20.7 | 2.3 |
| Survey 6 | NTR | 484 (67%) | 22.5 | 2.8 | 21.6 | 2.7 | 23.2 | 2.4 |
| Survey 7 | NTR | 501 (70%) | 23.3 | 3.2 | 21.9 | 2.9 | 25.4 | 2.5 |
| (*) bb1 - biobanking | NTR | 665 (69%) | 23.5 | 3.2 | 22.5 | 3.2 | 26.8 | 2.3 |
| Growth factors |  |  |  |  |  |  |  |  |
| Intercept | FinnTwin12 | 651 (59%) | 17.6 | 2.4 | 17.6 | 2.0 |  |  |
| Slope (non-linear) | FinnTwin12 | 651 (59%) | 2.3 | 0.6 | 2.0 | 0.6 |  |  |
| Intercept | NTR | 665 (69%) | 21.9 | 2.4 | 21.2 | 2.2 |  |  |
| Slope (linear) | NTR | 665 (69%) | 0.3 | 0.1 | 0.2 | 0.1 |  |  |

#### Netherlands Twin Register

The Netherlands Twin Register (NTR) is a population-based cohort which includes twins and multiples from the Netherlands [32]. Adult participants of the NTR have completed a survey on health and behavior every 2-3 years since 1991. In contrast to the FinnTwin12 cohort, where data collection is based on age, the NTR data collection in adult twins is independent of age; adult participants are invited to fill out a survey every ∼3 years. The blood samples for the omics experiments described in this study were collected in participants of the NTR biobank project (2004-2008)[33]. Venous blood samples were drawn between 7 and 10 AM after overnight fasting, and fertile women had their blood sample drawn in their pill-free week or on day 2–4 of their menstrual cycle. Information on BMI was obtained during the home visit for blood collection as part of the NTR Biobank project. Additionally, we calculated BMI from self-reported height and weight from NTR Survey 5 (2000), Survey 6 (2002), and Survey 7 (2004).

Transcriptomics data was available for 3369 individuals, of which we selected 965 participants for whom blood sampling was performed before age 30 (i.e., age at blood sampling < 30 years old). Of these participants, we retained those for whom BMI was available at the time of blood sampling as well as at least one other BMI measure at Survey 5, Survey 6, or Survey 7. The final NTR sample comprised 665 participants with 2 to 4 BMI measurements over a mean follow-up period of ∼6 years (Table 1). The sample included 359 monozygotic twins (141 complete pairs) and 306 dizygotic twins (103 complete pairs).

### Omics processing

#### Proteomics

Proteins from the plasma samples of FinnTwin12 participants were subjected to Liquid Chromatography-Electrospray Ionization-Mass Spectrometry (LC-ESI-MS/MS) as described previously [34]. First, proteins were precipitated with acetone and subjected to in-solution digestion according to the standard protocol of the Turku Proteomics Facility (Turku Proteomics Facility, Turku, Finland). After digestion, peptides were desalted using a 96-well Sep-Pak C18 plate (Waters), evaporated to dryness, and stored at −20°C. A commercial kit (High Select™ Top14 Abundant Protein Depletion Mini Spin Columns, cat. Number: A36370, ThermoScientific) was used to deplete the 14 most abundant proteins from plasma before the proteomic analysis. Samples were first analyzed by independent data acquisition LC-MS/MS using a Q Exactive HF mass spectrometer. Data were further analyzed using Spectronaut software and included local normalization of the data [35], as described elsewhere [34]. Raw matrix counts were log_2_-transformed and kit-depleted proteins were removed, including human serum albumin (HSA), albumin, IgG, IgA, IgM, IgD, IgE, kappa and lambda light chains, alpha-1 acid glycoprotein, alpha-1 antitrypsin, alpha-2 macroglobulin, apolipoprotein A1, fibrinogen, haptoglobin, and transferrin. None of the participants had scores greater or less than 5 standard deviations from the mean on the first two principal components (PCs) derived from principal component analysis (PCA), indicating none of the participants were identified as outliers. We excluded proteins with >10% missing values. Following the identification of 4 batches, imputation of missing values was performed (proportion of missing values in dataset: 0.86%) using the lowest observed value per batch. Corrections for batch effects were performed with Combat [36]. The final proteomic dataset comprised 439 proteins and protein abundances were scaled, such that one unit corresponded to one standard deviation (sd) with zero mean.

#### Transcriptomics

Details of pre-processing can be found in Jansen et al. [37]. In short, heparinized whole blood was transferred into PAXgene Blood RNA tubes (Qiagen, Valencia, Florida, USA) within 20 minutes of sampling, and stored at −30°C. Total RNA was extracted using the PAXgene Blood RNA MDx kit protocol in 96-well format with the BioRobot Universal System (Qiagen, Valencia, Florida, USA). RNA quality and quantity was assessed by Caliper AMS90 with HT DNA5K/RNA LabChips [37–39].

Samples were randomly assigned to plates and co-twins were randomized across plates to avoid bias in family correlation estimates. For cDNA synthesis, 50 ng of RNA was reverse transcribed and amplified in a plate format on a Biomek FX liquid handling robot (Beckman Coulter, Brea, California, USA) using Ovation Pico WTA reagents per the manufacturer’s protocol (NuGEN, San Carlos, California, USA). Products purified from single-primer isothermal amplification were then fragmented and labeled with biotin using Encore Biotin Module (NuGEN). Prior to hybridization, the labeled cDNA was analyzed using electrophoresis to verify the appropriate size distribution (Caliper AMS90, HT DNA 5K/RNA LabChip). Samples were hybridized to Affymetrix U219 array plates, and array hybridization, washing, staining and scanning were carried out per the manufacturer’s protocol (GeneTitan, Affymetrix, Santa Clara, California, USA).

Gene expression data were required to pass standard Affymetrix quality control metrics (Affymetrix expression console). Probes were removed when their location was uncertain or if their location intersected a polymorphic single nucleotide polymorphism (SNP). Expression values were obtained using robust multi-array average normalization implemented in Affymetrix Power Tools (v 1.12.0). We excluded samples that displayed an average Pearson correlation below 0.8 with the probe set expression values of other samples and samples with incorrect sex chromosome expression. In the analyses, we retained only probes for genes encoding proteins associated with BMI or BMI changes in FinnTwin12. We attempted to match protein-coding gene names, however, we failed to retrieve three protein-coding genes (*AMY2A*, *C4A*, and *CFHR1*) identified in FinnTwin12 from the NTR transcriptomic data. We focused on log_2_-transformed gene-level expression in our analyses, where we obtained gene-level expression by averaging probe level expression per gene, for each gene associated with protein-coding genes. The same analyses were also performed for log_2_ probe expression as supplementary analyses.

#### Metabolomics

Metabolites were quantified from EDTA plasma samples using high-throughput proton nuclear magnetic resonance spectroscopy (^1^H-NMR) on a laboratory setup that combines a Bruker AVANCE III 500 MHz and a Bruker AVANCE III HD 600 MHz spectrometer (Nightingale Health Ltd, Helsinki, Finland)[40,41]. In short, this method provides simultaneous quantification of routine lipids, lipoprotein subclass profiling with lipid concentrations within 14 subclasses, fatty acid composition, and various low-molecular weight metabolites, including amino acids, ketone bodies, and glycolysis-related metabolites in molar concentration units.

Of the 651 participants from the FinnTwin12 sample, 638 had metabolomic data available of which the preprocessing has been described elsewhere [42]. The data included 114 metabolites, of which 13 had missing values. None of the metabolites had more than 10% missing values, we therefore imputed all missing values with the lowest observed value of each metabolic biomarker. Besides LDL cholesterol as derived from the NMR platform, LDL cholesterol was quantified using the Friedewald equation [43]. Similar to the proteomic data, no outliers were detected on the first two PCs. The metabolite values were log_2_-transformed and scaled so that one unit corresponded to a change of one sd, with a mean of zero.

In the NTR, with the exception of six non-overlapping metabolites (see Additional file 3, Table S2), we only retained metabolites that were significantly associated with BMI trajectories in FinnTwin12. Description for the NTR metabolomics (NMR platform) has been supplied in detail elsewhere [44]. Missing values represented less than 3% of the total data points, and no metabolite had more than 20% missing values, thus imputation was performed by the lowest observed value for each metabolite. Data were log_2_-transformed, and a technical variable indicating batch was used as a covariate in all relevant analyses.

#### Polygenic risk scores

PRSs were used to assess whether genetic susceptibility could explain the association of protein or metabolite levels with BMI trajectories. In FinnTwin12, genotype data for most participants (645 of 651) were available and genotyping was performed using the Illumina Human CNV370-Duo chip. Processing details are available elsewhere [45]. We calculated three PRSs describing the genetic susceptibility to BMI [46], Waist to Hip Ratio adjusted with BMI (WHR)[47], and CAD [48]. After correcting the PRSs for population stratification [49], by regressing out the top ten genetic PCs, the PRSs were scaled to a mean of zero and unit variance.

Of the 665 NTR participants, 576 were genotyped using multiple platforms (see Additional file 4). For genotyped NTR participants, we calculated the PRS of BMI [46], using the infinitesimal prior (LDpred-inf) in Ldpred [50]. Detailed information on genotyping and PRS calculation in NTR participants is included in the supplementary material (see Additional file 4). The PRS was first corrected for population stratification [49] and for technical batch (i.e., platform) by regressing out the top ten genetic PCs and platform indicator, respectively, and then the PRS was scaled to have a mean of zero and unit variance.

### Statistical analysis

The primary analyses comprised two sequential steps: 1) calculating growth factors summarizing BMI trajectories and 2) performing association analyses of omics layers with BMI trajectories using mixed effects models (Figure 1). Together, these analyses allowed for the joint investigation of longitudinal BMI measures and omics data, associating proteomic, transcriptomic, metabolomic, and PRS data with BMI trajectories. Genetic levers underlying protein abundances and the associations between proteins and BMI trajectories were further explored by univariate and bivariate twin modeling. A multi-omics correlation network was then constructed to put the omics variables associated with changes in BMI into perspective.

### Trajectories computation

#### Latent Growth Curve Modeling

Latent Growth Curve Modeling (LGCM) was used to summarize the anthropometric trajectories into intercepts and slopes, corresponding to baseline measurements and rates of change, respectively. LGCM analyses were run with the *lavaan* package version *0.6-12* [51] in R version *4.1.2*. To accommodate the shape of the BMI trajectories, for which non-linearity in time is likely, we considered two modeling schemes, corresponding to linear and logarithmic dependence of time, respectively. When fitting the model, BMI measures were linearly adjusted for age differences within waves, with reference ages corresponding to the observed means per wave in the FinnTwin12 and NTR cohorts (Table 1). We tested for adjustment of the growth factors for sex, and further adjusted the model when the effect of sex was significant (*p*-value <0.05). Missing BMI values were allowed during model fitting by maximum likelihood. We calculated robust standard errors to correct for clustering in the data due to family relatedness.

We assessed the goodness of fit of the linear and logarithmic schemes using the Comparative Fit Index (CFI), the Tucker-Lewis Index (TLI), and the Root Mean Square Error of Approximation (RMSEA). A model with RMSEA less than 0.06, CFI greater than 0.95, and TLI greater than 0.95 was considered a good fit [52,53]. Whenever the linear modeling did not enable good model fit, the logarithmic scheme was preferred when it did enable a good model fit.

#### Sensitivity analyses

We carried out two sensitivity analyses. The first sensitivity analysis was performed in FinnTwin12, which consisted of repeating the analyses of the association between protein levels and baseline BMI or changes in BMI with another marker of adiposity: the triponderal mass index (TMI). TMI is defined as mass divided by height cubed, in contrast to BMI dividing mass by the square of height [54]. BMI is a good indicator of adiposity for most age groups, but TMI is sometimes preferred as an indicator in children and adolescents [54–56].

We performed the second sensitivity analysis in NTR. In NTR, surveys were collected regardless of the age of the participants, thus, each wave comprised all participants that were 18 years or older at the time of the survey. Because LGCM corrected for within-survey age differences, we used a second approach that did not correct for wave-level age differences, which were larger in NTR than in FinnTwin12 (Table 1). Instead of considering reference ages by which ages are corrected, as in the main analyses, we took each individual’s actual age at baseline as the first time point and then modeled changes in BMI from this individual’s baseline age. We then quantified the associations between these changes in BMI with gene expression.

#### Mixed-effect models

To quantify associations between the plasma proteomics data and BMI trajectories, we used mixed-effects models. Mixed-effects modeling involves using two types of effects, fixed and random, the former being constant across individuals and the latter varying between individuals. We tested associations between proteins and BMI at blood sampling and slope of BMI (i.e., BMI changes) in FinnTwin12. We also tested associations between proteins and intercept of BMI (i.e., baseline BMI) as the intercept of BMI was that of young adolescents, whereas the BMI at blood sampling was that of young adults. We included age at blood sampling, age squared at blood sampling, sex, and interactions between sex and age and age squared as covariates. BMI intercept was also included as a covariate to correct for baseline BMI differences when testing associations between proteins and changes in BMI. The model included two random effects whose main function was to correct for clustering in the data due to family relatedness: a binary indicator for zygosity and the family identifier. We tested the association between the coefficients of the fixed effects and each protein using a *t*-test with Satterthwaite approximation [57] and Bonferroni correction. We considered an association significant if the Bonferroni corrected *p*-value was less than 0.05 (i.e., raw *p*-value < 0.05/439=1.1 ×10^-4^). The same modeling was used for identification of metabolites associated with BMI trajectories (Bonferroni: raw *p*-value < 0.05/114=4.4 ×10^-4^) as well as to quantify associations between PRSs and proteins (Bonferroni: raw *p*-value < 0.05/439=1.1 ×10^-4^).

We repeated the same modeling in NTR, replacing proteomic variables with probe- or gene-level expression of protein-coding genes identified in FinnTwin12, or with metabolites. We added a technical covariate to correct for batches in the modeling between metabolites and BMI trajectories. Multiple correction was applied to the number of tests performed, with the associations of BMI and changes in BMI with omic variables (e.g., metabolite, gene) studied independently. We did not examine the associations between the omics layers and the BMI intercept, as BMI at baseline and BMI at blood sampling reflected both BMI at adulthood. The association between PRS of BMI and gene expression at the probe and gene level was also quantified.

#### Classical twin modeling

We used classical twin models (CTMs) to quantify the genetic and environmental contributions to protein abundances, and to decompose the associations between protein and BMI trajectories into genetic and environmental correlations. CTMs hinge on the comparison between MZ and DZ twins [58,59], since MZ twins share approximately 100% of the genomic sequence of their DNA, while DZ twins share, on average, only 50% of their segregating genes. By comparing the magnitude of correlations in DZ twin pairs (*rDZ*) to those in MZ twin pairs (*rMZ*), we can infer the presence of genetic factors. In univariate CTM, the variance (V) of a trait can be decomposed into several components: additive genetic (A), non-additive or dominant genetic (D), shared or common environmental (C), and nonshared or unique environmental (E). In classical twin modeling of pairs reared together, simultaneous quantification of the C and D components is not possible. A ratio *rDZ*/*rMZ* above or below 0.5 is therefore an indication that C or D, respectively, can be modeled. Since the quantification of D requires considerable statistical power, we have only estimated A and E, with C where appropriate. Heritability (*h^2^*) is defined as A/V, and the standardized coefficients *e^2^* and *c^2^* denote the part of the variance V explained by E and C, respectively [60,61].

We first fitted saturated models to obtain estimates of the twin correlations by maximum likelihood (ML), including sex and age at blood sampling as covariates. Analyses were conducted in R software using the *OpenMx* package, version 2.20.7 [62–65]. In univariate settings, proteins for which *rMZ*/*rDZ*<0.5 were fitted by AE models. Proteins for which *rMZ*/*rDZ*>0.5 were fitted with ACE models, which were compared with AE, CE, and E only. We used Akaike Information Criterion (AIC) to choose the best model for these proteins. Negative variance estimates were allowed [66]. Model comparison by AIC is available in the supplementary material (see Additional file 3, Table S3).

Similarly, we decomposed phenotypically significant correlations between proteins and BMI trajectories. Since twin correlations indicated an AE model for BMI at blood sampling and changes in BMI in the univariate framework, we decomposed their phenotypic correlations with proteins into genetic (*rA*) and non-shared environmental (*rE*) correlations in the bivariate framework. The bivariate twin analysis between changes in BMI and protein abundance included the BMI intercept as a covariate in addition to sex and age. Proteins modeled by CE rather than AE (i.e., having *h^2^*=0) were excluded from the bivariate modeling. Cross-twin cross-trait correlations can be found in the Supplementary Document (see Additional file 3, Table S4, Table S5).

#### Multi-omic network construction

A correlation network was constructed to provide a multi-omics framework of the mechanisms underlying BMI change in FinnTwin12, using the significant findings from the prior analyses. The network included the 14 proteins that were associated with the change in BMI and the 3 PRSs. Metabolites associated with the change in BMI were also included, but the set of 48 highly correlated lipoproteins (out of 53 metabolites) were reduced by PCA into 3 principal components (LIPO.PC1, LIPO.PC2, and LIPO.PC3), explaining 87% of the variance. This allowed for better visual representation of the connections between omics. Each of the 25 final entries was linearly regressed on BMI slope, BMI intercept, and BMI at blood sampling, as well as simple and quadratic age at blood sampling, sex, age by sex interaction, and quadratic age by sex interaction. Correlations between these 25 residual variables were used to construct the multi-omics network. We colored the edges according to the significance of the correlation assessed by Pearson’s correlation (i.e., raw *p*-value < 0.05 or Bonferroni-corrected *p*-value < 0.05) and the direction of the correlation (i.e., positive or negative). In NTR, we used a similar methodology to correlate metabolomics residual variables (3 lipoprotein PCs and 5 low molecular weight molecules) with gene expression (*N*=14).

## Results

### Longitudinal development of BMI in the two cohorts

In FinnTwin12, only the slope of BMI was associated with sex (intercept: *p*=0.70; slope: *p* < 0.001) and was therefore further adjusted by sex in the modeling. Logarithmic modeling achieved the best model fits (RMSEA: 0.05 [95% confidence interval (CI): 0.03,0.06], TLI=0.97, CTI=0.98) compared to linear modeling (RMSEA: 0.09 [95% Confidence Interval (IC): 0.07,0.11], TLI=0.93, CTI=0.94). We therefore kept logarithmic modeling for the calculation of growth factor values. The mean BMI intercept was 17.6 kg.m^-2^. Male and female participants gained an average of 6.2 and 5.0 BMI units, respectively, during the 10-year follow-up period (Table 1).

In NTR, both the intercept and the slope were sex adjusted (intercept: *p*=0.01; slope: *p*=0.02) and linear modeling provided the best model fits (RMSEA: 0.04 [95% CI: 0.02,0.06], TLI=0.99; CTI=0.99). Table 1 describes the mean intercepts and slopes. Only twelve NTR participants had a decrease in BMI over the course of follow-up, and overall, the change in BMI in NTR showed substantial weight gain at the population level (*z*-value= 3.3, *p*=0.001). During the 6-year follow-up, male and female NTR participants gained an average of 1.5 and 1.3 BMI units, respectively, (Table 1). Growth factor variance was significantly non-zero in both FinnTwin12 and NTR datasets (*z*-values>3.0, *p*<0.01), indicating significant inter-individual differences longitudinally.

### Proteins and BMI trajectories in FinnTwin12

#### Phenotypic associations

We examined associations between plasma protein abundance and BMI trajectories (i.e., BMI at blood sampling and slope of BMI) from mixed models using scaled BMI trajectory markers (Figure 1). A total of 66 proteins out of 439 proteins showed a significant association with BMI at blood sampling, of which 42 had a negative and 24 had a positive association with BMI (Table 2). Proteoglycan 4 had the strongest association with BMI (coefficient: 0.46; *p*=2.1E-34); an increase of one sd in the abundance of proteoglycan 4 was associated with an increase of almost a half of sd in BMI (Table 2).

**Table 2:** Mixed model-derived estimates of proteins significantly associated with body mass index at blood sampling in FinnTwin12 participants **Legend:** Protein estimates whose abundance were significantly associated with body mass index are shown if Bonferroni-corrected coefficient null-test p-values were less than 0.05. Mixed-effects models included as covariates: age at blood sampling, age squared, sex, and interactions of sex with age and age squared. Random effects included the zygosity indicator variable and family identifiers. Body mass index was z-scored; fixed effects estimates indicate effect on 1 sd change. se: standard error.

| Protein |  | Fixed effect coefficients |  |  | P-value |  |
| --- | --- | --- | --- | --- | --- | --- |
| description | UniProt ID | estimate | se | t-value | raw | bonferroni |
| Neural cell adhesion molecule L1-like protein | O00533 | -0.16 | 0.04 | -4.1 | 4.9E-05 | 2.2E-02 |
| ADAM DEC1 | O15204 | -0.15 | 0.03 | -4.2 | 3.0E-05 | 1.3E-02 |
| Coagulation factor XIII A chain | P00488 | -0.18 | 0.04 | -5.1 | 5.0E-07 | 2.2E-04 |
| Complement factor B | P00751 | 0.15 | 0.04 | 4.1 | 5.5E-05 | 2.4E-02 |
| Antithrombin-III | P01008 | -0.25 | 0.04 | -6.7 | 4.3E-11 | 1.9E-08 |
| Complement C3 | P01024 | 0.15 | 0.04 | 4.1 | 5.1E-05 | 2.3E-02 |
| Apolipoprotein A-II | P02652 | -0.15 | 0.04 | -4.1 | 4.7E-05 | 2.1E-02 |
| C-reactive protein | P02741 | 0.31 | 0.04 | 8.5 | 1.3E-16 | 5.6E-14 |
| Serum amyloid P-component | P02743 | 0.34 | 0.04 | 9.0 | 1.7E-18 | 7.6E-16 |
| Transthyretin | P02766 | -0.19 | 0.04 | -4.8 | 2.2E-06 | 9.5E-04 |
| Vitamin D-binding protein | P02774 | -0.15 | 0.04 | -4.0 | 7.8E-05 | 3.4E-02 |
| C4b-binding protein alpha chain | P04003 | 0.23 | 0.04 | 6.2 | 1.2E-09 | 5.4E-07 |
| Apolipoprotein B-100 | P04114 | 0.21 | 0.04 | 5.7 | 2.0E-08 | 8.6E-06 |
| Sex hormone-binding globulin | P04278 | -0.33 | 0.05 | -7.2 | 1.5E-12 | 6.6E-10 |
| Pancreatic alpha-amylase | P04746 | -0.20 | 0.04 | -5.6 | 3.4E-08 | 1.5E-05 |
| Insulin-like growth factor I | P05019 | -0.15 | 0.04 | -4.0 | 6.0E-05 | 2.6E-02 |
| Fructose-bisphosphate aldolase B | P05062 | 0.14 | 0.04 | 4.0 | 7.2E-05 | 3.2E-02 |
| Apolipoprotein D | P05090 | -0.20 | 0.04 | -5.3 | 1.3E-07 | 5.8E-05 |
| Protein S100-A8 | P05109 | 0.21 | 0.04 | 5.8 | 9.3E-09 | 4.1E-06 |
| Complement factor I | P05156 | 0.23 | 0.04 | 6.5 | 2.2E-10 | 9.8E-08 |
| Cholinesterase | P06276 | 0.21 | 0.04 | 5.0 | 9.0E-07 | 3.9E-04 |
| Gelsolin | P06396 | -0.18 | 0.04 | -4.5 | 6.9E-06 | 3.0E-03 |
| Protein S100-A9 | P06702 | 0.16 | 0.04 | 4.6 | 4.6E-06 | 2.0E-03 |
| Apolipoprotein A-IV | P06727 | -0.17 | 0.04 | -4.3 | 1.8E-05 | 8.0E-03 |
| Complement component C8 gamma chain | P07360 | 0.17 | 0.04 | 4.4 | 1.0E-05 | 4.6E-03 |
| Corticosteroid-binding globulin | P08185 | -0.24 | 0.04 | -5.9 | 6.2E-09 | 2.7E-06 |
| 4F2 cell-surface antigen heavy chain | P08195 | -0.16 | 0.04 | -4.2 | 3.5E-05 | 1.5E-02 |
| 72 kDa type IV collagenase | P08253 | -0.20 | 0.04 | -5.5 | 6.5E-08 | 2.8E-05 |
| Complement factor H | P08603 | 0.33 | 0.04 | 9.1 | 9.0E-19 | 4.0E-16 |
| Complement C4-A | P0C0L4 | 0.23 | 0.04 | 6.3 | 6.5E-10 | 2.9E-07 |
| Serum amyloid A-1 protein | P0DJI8 | 0.18 | 0.04 | 4.8 | 1.8E-06 | 8.0E-04 |
| Serum amyloid A-2 protein | P0DJI9 | 0.18 | 0.04 | 4.8 | 2.2E-06 | 9.7E-04 |
| Complement component C7 | P10643 | -0.28 | 0.04 | -7.5 | 2.3E-13 | 1.0E-10 |
| Mast/stem cell growth factor receptor Kit | P10721 | -0.18 | 0.04 | -4.8 | 1.6E-06 | 7.1E-04 |
| Neural cell adhesion molecule 1 | P13591 | -0.15 | 0.04 | -3.9 | 9.7E-05 | 4.3E-02 |
| Bone marrow proteoglycan | P13727 | -0.15 | 0.04 | -4.1 | 4.0E-05 | 1.8E-02 |
| Carboxypeptidase N catalytic chain | P15169 | -0.16 | 0.04 | -3.9 | 8.7E-05 | 3.8E-02 |
| Metalloproteinase inhibitor 2 | P16035 | -0.14 | 0.04 | -3.9 | 1.0E-04 | 4.6E-02 |
| Fumarylacetoacetase | P16930 | 0.20 | 0.03 | 5.8 | 1.4E-08 | 6.1E-06 |
| Endoglin | P17813 | -0.18 | 0.04 | -5.0 | 9.4E-07 | 4.1E-04 |
| Insulin-like growth factor-binding protein 2 | P18065 | -0.18 | 0.04 | -4.9 | 1.5E-06 | 6.5E-04 |
| Vascular cell adhesion protein 1 | P19320 | -0.19 | 0.04 | -5.0 | 7.5E-07 | 3.3E-04 |
| Inter-alpha-trypsin inhibitor heavy chain H2 | P19823 | -0.20 | 0.04 | -5.6 | 2.8E-08 | 1.2E-05 |
| Inter-alpha-trypsin inhibitor heavy chain H1 | P19827 | -0.18 | 0.04 | -4.8 | 2.1E-06 | 9.2E-04 |
| C4b-binding protein beta chain | P20851 | 0.18 | 0.04 | 4.7 | 3.9E-06 | 1.7E-03 |
| Fibulin-1 | P23142 | -0.18 | 0.04 | -4.9 | 1.2E-06 | 5.3E-04 |
| Properdin | P27918 | 0.20 | 0.04 | 5.3 | 1.7E-07 | 7.3E-05 |
| Transketolase | P29401 | 0.16 | 0.04 | 4.2 | 2.8E-05 | 1.2E-02 |
| Kallistatin | P29622 | -0.19 | 0.04 | -5.2 | 2.7E-07 | 1.2E-04 |
| Serum amyloid A-4 protein | P35542 | 0.19 | 0.04 | 5.2 | 2.2E-07 | 9.9E-05 |
| Voltage-dependent calcium channel subunit alpha-2/delta-1 | P54289 | -0.16 | 0.04 | -4.4 | 1.6E-05 | 6.8E-03 |
| Cadherin-13 | P55290 | -0.17 | 0.04 | -4.4 | 1.1E-05 | 4.6E-03 |
| Basement membrane-specific heparan sulfate proteoglycan core protein | P98160 | -0.16 | 0.04 | -4.3 | 2.2E-05 | 9.8E-03 |
| Complement factor H-related protein 1 | Q03591 | 0.15 | 0.04 | 4.1 | 4.0E-05 | 1.7E-02 |
| Prolow-density lipoprotein receptor-related protein 1 | Q07954 | -0.15 | 0.04 | -4.0 | 6.6E-05 | 2.9E-02 |
| Contactin-1 | Q12860 | -0.20 | 0.04 | -5.4 | 8.5E-08 | 3.7E-05 |
| Secreted phosphoprotein 24 | Q13103 | -0.30 | 0.04 | -8.1 | 3.4E-15 | 1.5E-12 |
| Limbic system-associated membrane protein | Q13449 | -0.19 | 0.04 | -5.2 | 3.0E-07 | 1.3E-04 |
| SPARC-like protein 1 | Q14515 | -0.15 | 0.04 | -4.2 | 3.6E-05 | 1.6E-02 |
| Serum paraoxonase/lactonase 3 | Q15166 | -0.16 | 0.04 | -4.3 | 2.2E-05 | 9.4E-03 |
| Transforming growth factor-beta-induced protein ig-h3 | Q15582 | -0.17 | 0.04 | -4.6 | 6.1E-06 | 2.7E-03 |
| Adiponectin | Q15848 | -0.15 | 0.04 | -4.1 | 4.7E-05 | 2.1E-02 |
| ADAMTS-like protein 4 | Q6UY14 | -0.19 | 0.04 | -5.2 | 2.7E-07 | 1.2E-04 |
| Gamma-glutamyl hydrolase | Q92820 | 0.15 | 0.04 | 4.2 | 3.1E-05 | 1.4E-02 |
| Proteoglycan 4 | Q92954 | 0.46 | 0.03 | 13.6 | 4.7E-37 | 2.1E-34 |
| Interleukin-1 receptor accessory protein | Q9NPH3 | -0.20 | 0.04 | -5.4 | 1.1E-07 | 4.7E-05 |

A total of 14 protein abundances were associated with changes in BMI, independent of baseline BMI levels (Table 3). These proteins included complement factors and proteoglycan 4, with proteoglycan 4 having the strongest association with adolescent BMI changes (estimate: 0.27; *p*=4.3E-10). Ten proteins had a positive association with changes in BMI and 4 had negative associations. Thirteen out of 14 proteins were also correlated with BMI at blood sampling.

**Table 3:** Mixed model-derived estimates of proteins significantly associated with the changes (slope) of body mass index over 10 years of follow-up in FinnTwin12 participants **Legend:** Protein estimates whose abundance were significantly associated with changes in body mass index (i.e., slope of body mass index) are shown if Bonferroni-corrected coefficient null-test p-values were less than 0.05. Mixed-effects models included as covariates: the intercept of body mass index, age at blood sampling, age squared, sex, and interactions of sex with age and age squared. Random effects included the zygosity indicator variable and family identifiers. The slope of body mass index was z-scored; fixed effects estimates indicate effect on 1 sd change. se: standard error.

| Protein |  | Fixed effect coefficients |  |  | P-value |  |
| --- | --- | --- | --- | --- | --- | --- |
| description | UniProt ID | estimate | se | t-value | raw | bonferroni |
| Complement factor B | P00751 | 0.16 | 0.04 | 4.3 | 1.9E-05 | 8.1E-03 |
| C-reactive protein | P02741 | 0.16 | 0.04 | 4.2 | 2.7E-05 | 1.2E-02 |
| Serum amyloid P-component | P02743 | 0.18 | 0.04 | 4.6 | 6.4E-06 | 2.8E-03 |
| C4b-binding protein alpha chain | P04003 | 0.18 | 0.04 | 4.8 | 1.7E-06 | 7.6E-04 |
| Phosphatidylcholine-sterol acyltransferase | P04180 | 0.16 | 0.04 | 4.3 | 2.3E-05 | 9.9E-03 |
| Sex hormone-binding globulin | P04278 | -0.24 | 0.05 | -5.1 | 4.5E-07 | 2.0E-04 |
| Complement factor I | P05156 | 0.16 | 0.04 | 4.5 | 1.0E-05 | 4.4E-03 |
| Cholinesterase | P06276 | 0.24 | 0.04 | 5.6 | 3.0E-08 | 1.3E-05 |
| Complement factor H | P08603 | 0.25 | 0.04 | 6.6 | 7.7E-11 | 3.4E-08 |
| Complement component C7 | P10643 | -0.20 | 0.04 | -5.2 | 2.4E-07 | 1.1E-04 |
| Inter-alpha-trypsin inhibitor heavy chain H2 | P19823 | -0.15 | 0.04 | -4.0 | 8.1E-05 | 3.6E-02 |
| Properdin | P27918 | 0.15 | 0.04 | 4.0 | 7.1E-05 | 3.1E-02 |
| Secreted phosphoprotein 24 | Q13103 | -0.17 | 0.04 | -4.3 | 2.2E-05 | 9.9E-03 |
| Proteoglycan 4 | Q92954 | 0.27 | 0.04 | 7.3 | 9.8E-13 | 4.3E-10 |

Similarly, we investigated associations between proteins and BMI intercept (i.e., baseline BMI; see Additional file 3, Table S6), of which 11 significant associations were detected. Of these, 9 were also associated with BMI at blood sampling. These findings suggest that the proteome in young adults contains proteins associated with both childhood and adulthood BMI. Proteoglycan 4, complement factor H, C-reactive protein, and secreted phosphoprotein 24 were associated with BMI at blood sample collection, BMI intercept, and BMI slope.

Associations between proteins with triponderal mass index (TMI) at baseline (∼12 years old) and slope of TMI can be found in the supplementary material (see Additional file 3, Table S7). In total, we found 10 and 22 proteins associated with baseline TMI and TMI changes, respectively. Seven of the ten proteins associated with baseline TMI were also associated with baseline BMI. All proteins associated with BMI changes were also associated with TMI changes. This shows that BMI and TMI associations with the plasma proteome tend to have similar proteins involved.

#### Genetic and environmental sources underlying protein abundance

Univariate twin modeling was used to quantify the genetic and environmental sources of variance in the 66 proteins identified as associated with either BMI at blood sampling and/or the slope of BMI (Figure 2A). According to the ratio between *rMZ* and *rDZ* (Figure 2B), we obtained 13 CE models, 1 ACE model, and 52 AE models. On average, genetic and nonshared environmental factors explained 35% (range: 0–78) and 59% (range: 22–89) of the variance in protein abundances, respectively (Figure 2C). Thirteen proteins showed no evidence for genetic effects (*h^2^*=0), and all of these had non-zero shared environmental components that accounted for an average of 28% (range: 19-41) of the variance, with nonshared environmental factors accounting for 72% (range: 59-81) of the variance in the abundances of these proteins (Figure 2A). Heritability of BMI at blood sampling was 72% [95% CI: 62,78], and the nonshared environment accounted for the remaining (28%) of the variance in BMI at blood sampling. Heritability of the slope of BMI was 63% [95% CI: 51,73] while the nonshared environment explained the remaining (37%) variance in BMI changes. All univariate estimates and their 95% CIs are available in the supplementary material (see Additional file 3, Table S3, Table S8).

**Figure 2:**
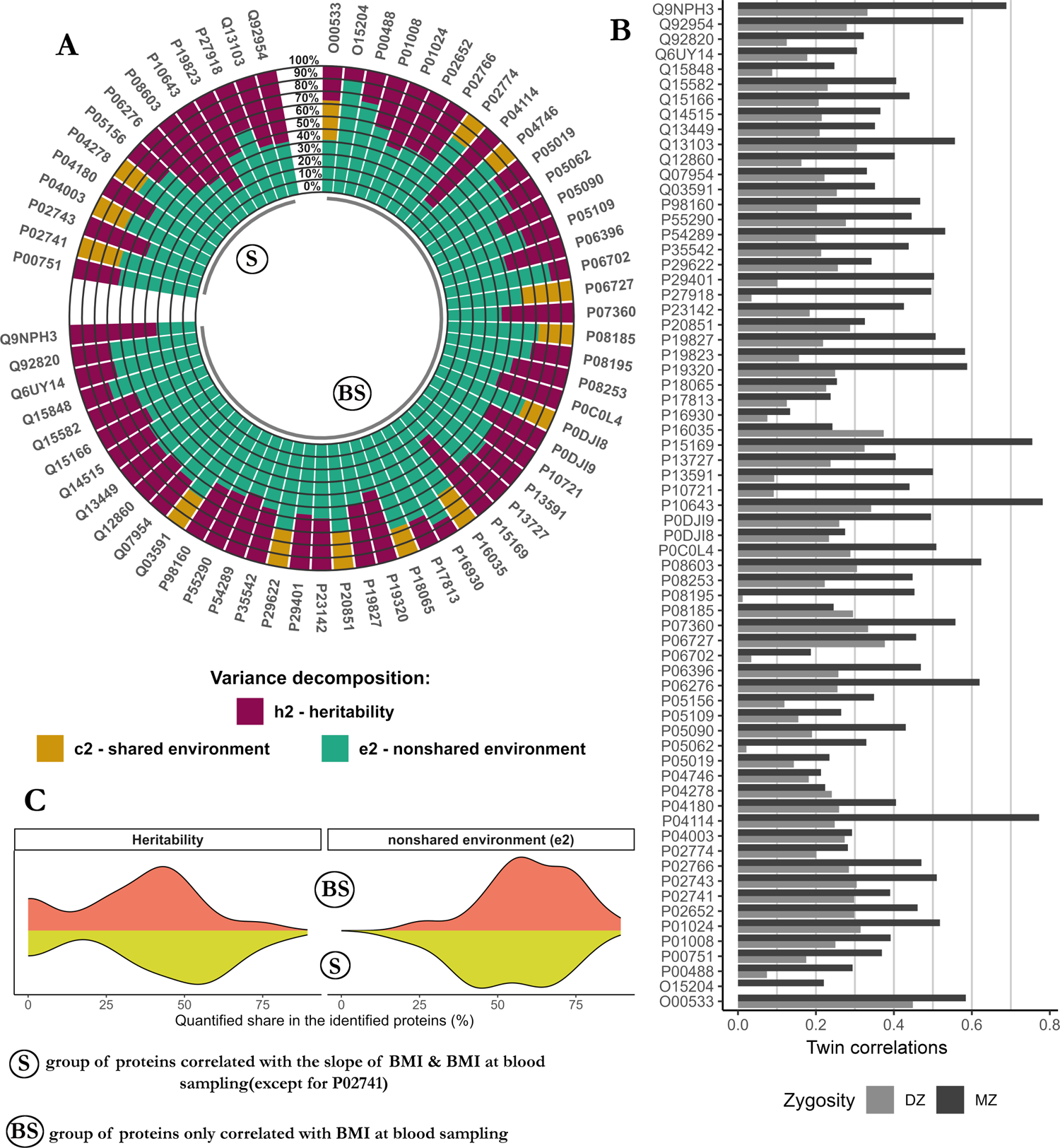
Genetic and environmental sources underlying proteins for which plasma abundance was associated with BMI or BMI change in the FinnTwin12 sample **legend:** (A) Univariate twin models were used to decompose the variance of proteins identified as associated with BMI or BMI changes in FinnTwin12. (B) The models were determined by the ratio of the correlations in MZ and the correlations in DZ twins. (C) Fractions attributed to non-shared environmental factors and to heritability. BMI: body mass index.

#### Genetic and environmental correlations between BMI trajectories and proteins

The decomposition of significant phenotypic covariation between plasma protein abundances (*h^2^*>0) with BMI and BMI changes were performed with bivariate genetic twin modeling. For all associations between proteins and BMI at blood sampling or BMI change, we applied bivariate AE models. Of the 53 BMI-protein correlations tested, we observed 43 significant genetic correlations and 12 significant nonshared environmental correlations (Figure 3A). For 8 protein–BMI associations, we observed both significant genetic and nonshared environmental correlations. Overall, genetic correlations ranged from −0.50 to 0.50 (absolute mean: *rA*=0.29) and environmental correlations from −0.33 to 0.44 (absolute mean: *rE*=0.14). Seven proteins had genetic correlations with BMI greater than 0.4 in absolute value, and proteoglycan 4 had the highest nonshared environmental correlation with BMI among all proteins tested (*rE*= 0.44 [95% CI: 0.28,0.57]). The full set of genetic and nonshared environmental correlations is available in the supplemental material (see Additional file 3, Table S9).

**Figure 3:**
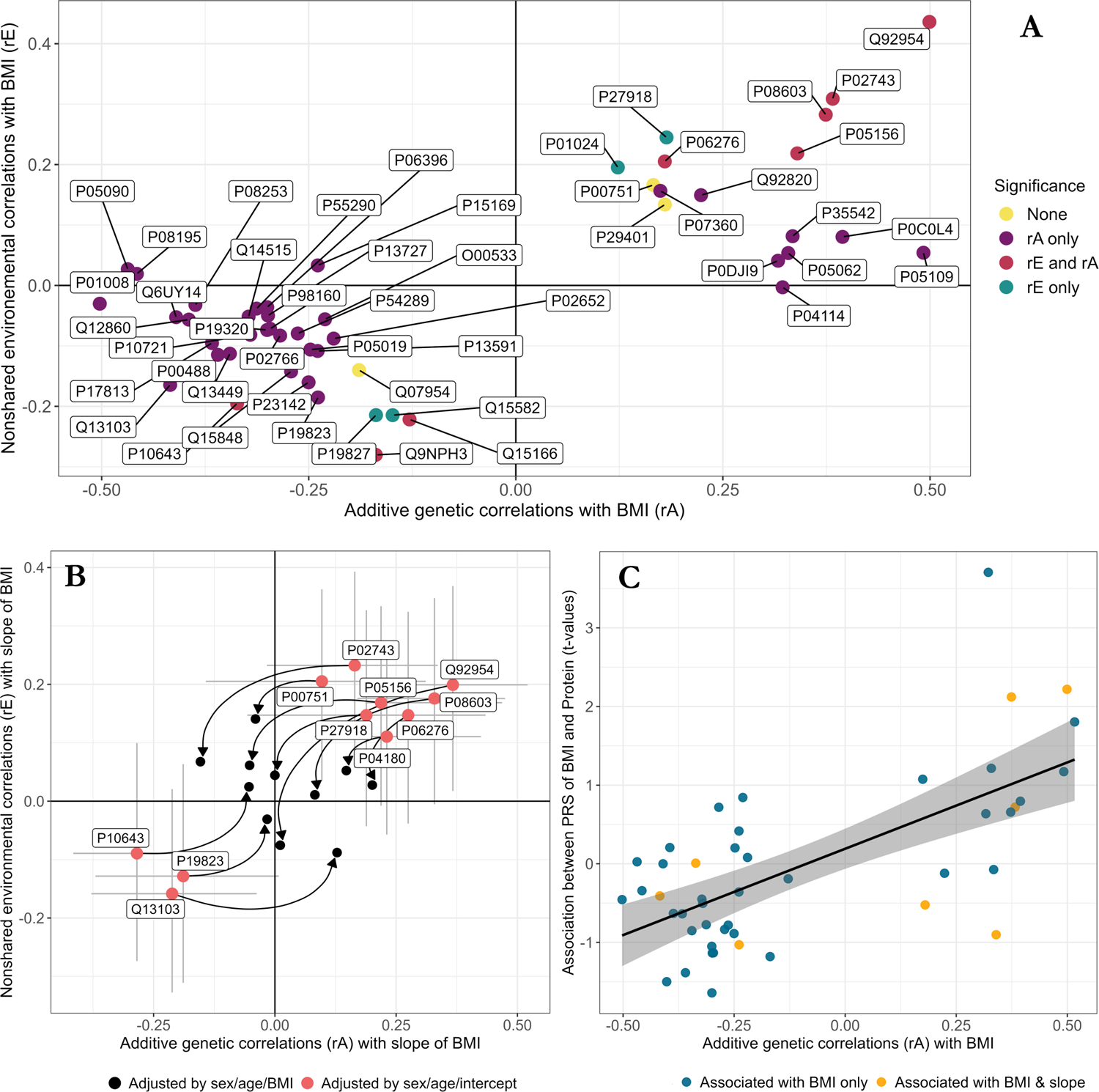
Genetic and non-shared environmental correlations between BMI trajectories and protein levels in the FinnTwin12 sample **legend:** (A) Among the 53 heritable proteins for which associations with BMI were significant, bivariate twin models were conducted to dissect the nature of the associations. (B) Genetic and non-shared environmental correlations were also quantified in the association between slope of BMI and protein levels; lines correspond to confidence intervals at the 95% threshold, arrows point to estimates obtained by adjusting protein levels by BMI at blood sampling prior to bivariate twin modeling rather than using BMI intercept as a covariate (pink). (C) The direction of the genetic correlations observed between BMI at blood sampling and protein levels were consistent with the t-values observed by crossing the PRS of BMI with protein levels. BMI: body mass index. PRS: polygenic risk score.

Similarly, the significant associations between proteins and changes in BMI were decomposed into genetic and nonshared environmental correlations for 11 of the 14 proteins associated with changes in BMI (Figure 3B). We observed 6 significant genetic correlations and 4 nonshared environmental correlations (see Additional file 3, Table S10). Because BMI change during adolescence predicted adult BMI independently of baseline BMI (linear regression: *R^2^*=45%), we examined whether the association between BMI at blood sampling and BMI change might confound the observed correlations between proteins and BMI. After adjusting the slope of BMI by BMI at blood sampling before bivariate twin modeling (Figure 3B), only apolipoprotein A-IV remained genetically correlated with the slope of BMI (*rA*= 0.20 [95% CI: 0.02, 0.38]), while all other correlations were no longer significant (see Additional file 3, Table S11). These results suggest that the genetic and environmental factors influencing the association between changes in BMI during adolescence and protein levels in adulthood may reflect associations with adult BMI.

### Gene expression and BMI trajectories in NTR

Mixed-effects models were used to quantify the associations of BMI trajectories with the expression of genes encoding the proteins identified in FinnTwin12. Only *S100A8* (protein encoded: S100 Calcium Binding Protein A8) was significantly associated with BMI at blood sampling (estimate=0.21; sd=0.05; *p*=1.5E-03). The *PRG4* gene (protein encoded: proteoglycan 4) was significantly associated with the slope of BMI (estimate=0.09; sd=0.03; *p*=0.05). While the association of the *CFI* gene (protein encoded: complement factor I) with the slope of BMI did not pass Bonferroni correction (*p*=0.21), one of its probes (ID: 11718481_s_at; start: chr4:110661852; end: chr4:110663751) was significantly associated with changes in BMI (*p*=0.02) at the probe level. Summary statistics are available in the supplementary material (see Additional file 3, Table S12, Table S13, Table S14, Table S15), and estimates from the sensitivity analysis are also available in the supplementary material (see Additional file 3, Table S16, Table S17).

### Metabolites and BMI trajectories

A total of 78 out of 114 metabolites were significantly associated with BMI at blood sampling in FinnTwin12. The mean absolute effect, which is 1.4 times greater than that observed for proteins, was 0.28 sd change in BMI per 1 sd change in metabolite level. Of the 78 metabolites, 68 were lipoproteins. The rest included Friedewald LDL cholesterol [43] and 9 low molecular weight molecules (LMWM): citrate, glycerol, glycoprotein acetyls, isoleucine, leucine, valine, CH2 groups of mobile lipids, phenylalanine, and tyrosine. In NTR, 44 out of the 78 metabolites were also significantly associated with BMI at blood sampling (Figure 4A; see Additional file 3, Table S18, Table S2).

**Figure 4:**
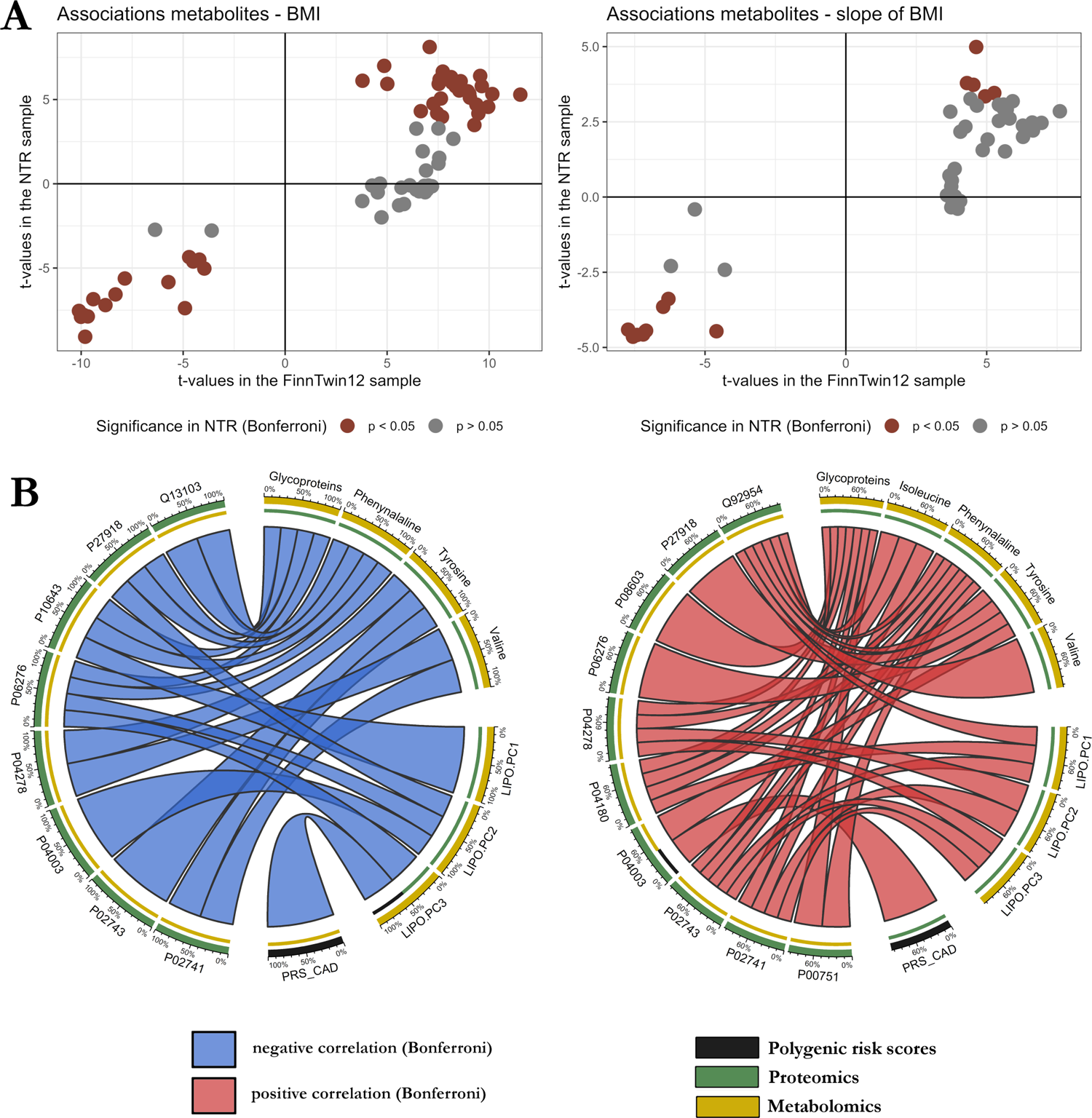
Extended cross-omic analyses **legend:** (A) The t-values of the associations between metabolites with BMI and BMI change have proportional directions between the two samples NTR and FinnTwin12. (B) Multi-omic correlations between PRSs with proteins and metabolites associated with BMI change in the FinnTwin12 sample, independent of BMI trajectory markers (BMI blood sampling, BMI slope and BMI intercept). PRS: polygenic risk score. BMI: body mass index. NTR: Netherlands Twin Register.

In FinnTwin12, 53 metabolites were associated with the slope of BMI, and all of these were also associated with BMI at the time of blood sampling. These metabolites comprised 48 lipoproteins and five LMWMs: glycoprotein acetyls, isoleucine, valine, phenylalanine, and tyrosine. In NTR, 14 of the 53 metabolites were also significantly associated with the BMI slope (Figure 4A), of which 13 were lipoproteins besides glycoprotein acetyls. In FinnTwin12, 19 metabolites were associated with the intercept of BMI (i.e., baseline BMI; baseline age: ∼12 years). Summary statistics are available in the supplementary material (see Additional file 3, Table S19, Table S20, Table S21).

### Polygenic risk scores and BMI-associated omics traits

We first examined the association of PRSs with proteins identified as associated with BMI and changes in BMI in FinnTwin12. Of the 66 proteins associated with BMI at blood sampling, none were associated with the PRS of BMI after Bonferroni correction (*p* > 0.05; see Additional file 3, Table S22). However, we observed consistent directions of effect between the genetic correlations of proteins and BMI and the associations between the PRS of BMI and proteins (Figure 3C). In NTR, we quantified the association of BMI protein-encoding gene expression with the PRS of BMI, but found no significant associations. We then compared the *t*-values of associations between gene expression and the PRS of BMI with the *t*-values of associations between gene expression and BMI, and found weak evidence for consistency in the direction of the compared *t*-values (linear regression coefficient nullity-test: *p*=0.015).

In FinnTwin12, we also investigated the associations between proteins and the PRSs of CAD and WHR (see Additional file 3, Table S22). None of the associations between proteins and the PRS of WHR were significant, with P06702 (Bonferroni-corrected *p*-value=0.06) and Q15848 (*p*=0.10) having the lowest Bonferroni-corrected *p*-values. We found two significant associations between proteins and the PRS of CAD: C4b-binding protein alpha chain (protein ID: P04003; raw *p*-value=6.0E-04; Bonferroni *p*-value=4.0E-02) and Apolipoprotein B-100 (protein ID:P04114; raw *p*-value=5.2E-04; Bonferroni *p*-value=3.4E-02).

We also conducted additional analyses between PRSs and proteins associated with the slope of BMI in the FinnTwin12 cohort (see Additional file 3, Table S25), between the PRS of BMI and genes encoding the proteins identified associated with the slope of BMI in NTR (for gene level results see Additional file 3, Table S26, and for probe level results see Additional file 3, Table S27), and between PRSs and metabolites in both cohorts (see Additional file 3, Table S28, Table S29).

### Multi-omic correlation network

To examine the association of the proteome, metabolome, and PRSs with BMI trajectories, we constructed a multi-omics correlation network in FinnTwin12 (Figure 4B) with previously identified proteins and metabolites adjusted for covariates and BMI at blood sampling, slope of BMI, and BMI intercept. We observed 49 correlations between omic layers, of which 30 were positive and 19 negative (Figure 4B). Apart from associations between the PRS of CAD with P04003 and LIPO.PC3, the remainder were metabolome-proteome associations. Twelve of the fourteen proteins associated with changes in BMI shared at least one significant correlation with a residual metabolomic variable, the average number of connections being 3.9 out of 8 possible connections for these proteins. These correlations indicate multi-omic relationships between omics variables associated with a 10-year change in BMI, independent of BMI trajectories markers. We also examined the associations between the transcriptome and metabolome in NTR, but found no significant multi-omics relationships (see Additional file 3, Table S30, Table S31).

## Discussion

Overall, a substantial number of plasma proteins were associated with adult BMI and BMI change during adolescence. The estimates for the heritability of plasma abundance of the identified proteins varied. For some proteins, familial resemblance was influenced by shared (i.e., familial) environmental factors rather than genetics. The etiology of the observed protein-BMI trajectory associations revealed genetic and/or nonshared environmental correlations between BMI and protein levels. Few associations between blood expression levels of protein-encoding genes and BMI trajectories were observed in NTR adults. This suggests a potential for observing gene-protein pairs associated with BMI trajectories at two different omic levels, among both adolescents and adults from two different countries. Finally, the use of metabolomic data and PRSs allowed both the observation of cross-omic associations and a better understanding of the connections that link BMI trajectories to different omic layers.

The twin design allowed the quantification of the genetic and environmental sources of variation underlying the associations between BMI trajectories and the proteome from an epidemiologic perspective. Including two cohorts of different ages and populations also empowered the identification of biomolecules likely to be stable across generations and populations. This is in line with two longitudinal multi-omics studies that identified stable biomolecules in response to interventions and investigated proteome resistance to weight change [18,19]. Although BMI measurements at blood sampling were similar between the two cohorts (Table 1), both the length of follow-up and the intensity of BMI changes between the two cohorts differed. This may have limited our ability to compare biomolecule associations with changes in BMI between cohorts. Moreover, the relatively small sample sizes of the cohorts in the current study might have limited the statistical power to detect associations with PRSs or transcriptomic data. Using less conservative multiple testing corrections and cohorts with larger sample sizes may allow more associations to be detected. However, availability of larger multi-omics twin data is limited. An increase in the size and number of omics data in twin cohorts is therefore necessary to realize the full potential of twin designs.

BMI changes in FinnTwin12 adolescents were substantial from early adolescence to young adulthood in both male and female participants. These changes reflect multiple processes such as increase in lean body mass, and increase in body fat as growth continued from prepuberty onwards. Longitudinal omics data would allow for a more in-depth investigation of the relationship that links changes in BMI to the proteome or metabolome. Investigations among adolescents need to evaluate changes in body composition, while among adults BMI weight gain is primarily change in fat depots. Beyond describing body composition, it is still needed to characterize the causal relationships between changes in BMI and proteins or metabolites. Twin designs combining MR and direction of causation (DOC) twin models, e.g. MR-DOC models, could be used for this purpose [67]. However, there is a lack of sufficiently large genome-wide association studies of BMI changes, which precludes the use of sufficiently robust instrumental variables.

Complement factors associated with BMI at blood sampling in FinnTwin12, including complements I, B, and H, have been associated with BMI previously [11,12,68]. Other proteins with strong associations with BMI at blood sampling (|estimate|>0.3; Table 2) in FinnTwin12 replicate findings from the literature, such as C-reactive protein [11,12,18,68], sex hormone-binding globulin [11,12,68], and proteoglycan 4 [69]. Several proteins associated with BMI at blood sampling have also been reported in the literature to be causally associated with BMI. These include those encoded by the *CRP*, *NCAM1*, *IGFBP2*, or *SHBG* genes [11,12], the last four of which are causally influenced by BMI (i.e., BMI-to-protein association) [12].

We identified several of the proteins associated with changes in BMI (i.e., gains in BMI) during adolescence that were also identified in adult populations that experienced weight loss. For example, proteoglycan 4, which was most strongly associated with BMI changes in our study, was previously associated with weight loss in adults [25] with the same direction of effect, i.e., positive covariation between change in BMI and proteoglycan 4 level. C-reactive protein, although previously shown to rapidly respond to weight loss [25], was also associated with changes in BMI over a 10-year follow-up in our study. These observations suggest the proteins identified in adult populations are, at least in part, relevant to the study of adolescents. Moreover, we observed that the direction of the effect between protein levels and BMI change echoed those in the literature, i.e., positive associations with BMI gain were negative with weight loss and vice versa. This indicates a stability of certain biomolecules in the proteome to characterize changes in BMI across ages, both in the study of weight gain and weight loss. Weight loss involves changes in lean mass as well as in body fat [70], which would be analogously the reverse regarding growth in adolescence.

Overall, we found few proteins for which the expression of their coding genes were associated with BMI or changes in BMI. Post-transcriptional modifications affecting proteins, e.g., the post-transcription addition of chemical groups or polypeptides [71,72], can explain this lack of association, as they increase the complexity of the transcriptome–proteome connection. However, we observed that the gene expression of *S100A8* (protein: S100A8), *PRG4* (protein: proteoglycan 4), and *CFI* (protein: complement factor I) were associated with BMI at blood sampling, BMI changes at the gene level, and BMI changes at the probe level, respectively. The *S100A8* gene encodes the eponymous protein belonging to the S100 family corresponding to Ca2+-binding proteins widely known to be associated with BMI [73] in both adults [74,75] and children [76]. *S100A8* expression levels have previously been associated with BMI [77], echoing the dual association with BMI at both the transcriptome and proteome levels in our study. In contrast, no previous study reported associations between the gene expression of *CFI* and *PRG4* genes with changes in BMI to our knowledge.

Heritability estimates of BMI trajectories in FinnTwin12 adolescents (*h^2^*=63%) were slightly larger than those observed in adult Finnish twins (*h^2^*= 52–57%) [78], which is consistent with the higher heritability of BMI reported in younger populations [6] and further motivates the study of factors genetically associated with BMI changes during adolescence. Of body mass, fat accounts for a smaller proportion in children and adolescents in general, while gaining fat is more typical in adult populations The previous estimates for the heritability of plasma protein abundance associated with BMI trajectories appear to be rare in the literature. Liu et al. reported that the heritability of 342 plasma proteins in a relatively small sample of older twins [79] varied widely, which is in line with our observations. Other studies have quantified the heritability of disease-associated plasma proteins [80], but none seem to focus on obesity. The heritability estimates of the proteins identified in our study therefore constitute a valuable resource for the further study of the proteome–obesity relationship.

C4b-binding protein alpha chain (Bonferroni *p*-value=4.0E-02) and apolipoprotein B-100 (*p*=3.4E-02) were associated with the PRS of CAD, with apolipoprotein B-100 being genetically correlated to BMI (*rA*=0.32). While the links between C4b-binding protein alpha chain and CAD are new, apolipoprotein B-100 is genetically associated with cardiovascular disease [81,82], as well as chronic kidney disease, blood pressure, and various lipids [83,84]. A better understanding of the mechanisms induced or inferred by this protein could provide insights into both obesity and its co-morbidities.

We identified many metabolites associated with BMI and BMI changes in FinnTwin12, of which we replicated a substantial proportion in NTR. These metabolites included mostly lipoproteins and branched-chain amino acids with known associations with BMI changes [27]. Despite the strong connections observed between the metabolome and proteome in the study of BMI trajectories, we found no significant connections between the metabolome and the transcriptome. Despite the lack of results, which could be explained by statistical power issues, it is still unclear how the different omics layers are connected biologically. Having omics from multiple tissues would be one approach, but perhaps feasible only in model organisms. The identification of mediation mechanisms between omics layers may help in acquiring a better understanding of the multi-omics mechanisms underlying obesity [85,86].

## Conclusions

In conclusion, the proteome is a promising resource for cross-sectional and longitudinal studies of obesity. Both environmental and genetic factors influence the proteome–obesity association, and integrating multiple omics may pave the way for a better understanding of the biological mechanisms underlying obesity. We observed proteins associated with weight gain in adolescents that are also associated with weight loss in adults in the literature, suggesting that these may have more to do with increases and decreases of lean mass, respectively, than changes in body fat. Future studies with different designs (i.e., different populations, ages, or follow-up lengths) could provide a holistic view of how the proteome-obesity relationship is expressed.

## List of abbreviations

BMI: Body mass index

PRSs: polygenic risk scores

CAD: coronary artery disease

NTR: Netherlands Twin Register

HSA: human serum albumin

PCs: principal components

PCA: principal component analysis

sd: standard deviation

SNP: single nucleotide polymorphism

^1^H-NMR: high-throughput proton nuclear magnetic resonance spectroscopy

LGCM: Latent Growth Curve Modeling

CFI: Comparative Fit Index

TLI: Tucker-Lewis Index

RMSEA: Root Mean Square Error of Approximation

TMI: triponderal mass index

CTMs: classical twin models

*rDZ*: correlations in DZ twins

*rMZ*: correlations in MZ twins

V: variance

A: additive genetic component

D: non-additive or dominant genetic component

C: shared or common environmental component

E: nonshared or unique environmental component

*h^2^*: heritability

ML: maximum likelihood

AIC: Akaike Information Criterion

*rA*: genetic

*rE*: nonshared environmental

CI: correlations; confidence interval

LMWM: low molecular weight molecules

DOC: direction of causation.

## Declarations

### Ethics approval and consent to participate

In FinnTwin12, ethical approval for all data collection waves was obtained from the ethical committee of the Helsinki and Uusimaa University Hospital District and the Institutional Review Board of Indiana University. All data collection and sampling protocols were performed in compliance with the ethical guidelines. Parents provided consent for the twins aged 12 and 14 years old, while twins aged 17 and 22 years old provided written consent themselves for sample collection.

In NTR, informed consent was obtained from all participants. Projects were approved by the Central Ethics Committee on Research Involving Human Subjects of the VU University Medical Centre, Amsterdam, an Institutional Review Board certified by the U.S. Office of Human Research Protections (IRB number IRB00002991 under Federal-wide Assurance-FWA00017598; IRB/institute code, NTR 03–180).

### Consent for publication

Not applicable.

### Availability of data and materials

FinnTwin12 data analyzed in this study is not publicly available due to the restrictions of informed consent. Requests to access these datasets should be directed to the Institute for Molecular Medicine Finland (FIMM) Data Access Committee (DAC) for authorized researchers who have IRB/ethics approval and an institutionally approved study plan. To ensure the protection of privacy and compliance with national data protection legislation, a data use/transfer agreement is needed, the content and specific clauses of which will depend on the nature of the requested data.

The data of the Netherlands Twin Register (NTR) may be accessed, upon approval of the data access committee, through the NTR (https://tweelingenregister.vu.nl/information_for_researchers/working-with-ntr-data).

## Authors’ contributions

The study design was developed and discussed by GD, FAH, AW, NH, SR, MP, DIB, JD, and JK. The data were collected by GW, EJCG, KK, DIB, JD, and JK, and their availability for analysis was facilitated by RP and AF. Processing of FinnTwin12 data was performed by GD and AW. NTR data were processed by GD, FAH, RP, JH, and RJ. The bioinformatic and statistical analyses were performed by GD. GD and FAH wrote the original manuscript. All authors actively participated in the improvement of the manuscript by critically revising it. All of the authors read and approved the final version of the manuscript.

## Data Availability

FinnTwin12 data analyzed in this study is not publicly available due to the restrictions of informed consent. Requests to access these datasets should be directed to the Institute for Molecular Medicine Finland (FIMM) Data Access Committee (DAC) for authorized researchers who have IRB/ethics approval and an institutionally approved study plan. To ensure the protection of privacy and compliance with national data protection legislation, a data use/transfer agreement is needed, the content and specific clauses of which will depend on the nature of the requested data. The data of the Netherlands Twin Register (NTR) may be accessed, upon approval of the data access committee, through the NTR (https://tweelingenregister.vu.nl/information_for_researchers/working-with-ntr-data).

## Acknowledgements

We gratefully acknowledge the contribution of the Turku Proteomics Facility team supported by Biocenter Finland for mass spectrometry. We warmly thank the participants involved in this study.

