## Additional file 2 for "Longitudinal multi-omics study reveals common etiology underlying association between plasma proteome and BMI trajectories in adolescent and young adult twins"

### Supplementary document

*Drouard et al.*

last update: April 2023

### 1 Validity of self-reported *versus* measured anthropometric measurements

We wished to investigate the validity of self-reported weight and height measurements in FinnTwin12. Of the 786 FinnTwin12 participants who had blood sampling, 756 had a clinical measure of their weight (kg) and height (cm) in addition to their self-reported weights (kg) and heights (cm). Five median days separated clinical measurements from receipt of self-reported measurements. Thirteen days at most separated weight and height measurements from self-reported measurements for 95% of these participants. Four participants had reported their weight and height more than 1 year before their clinical measurement, and were therefore excluded to improve the assessment of the validity of self-reported measurements.

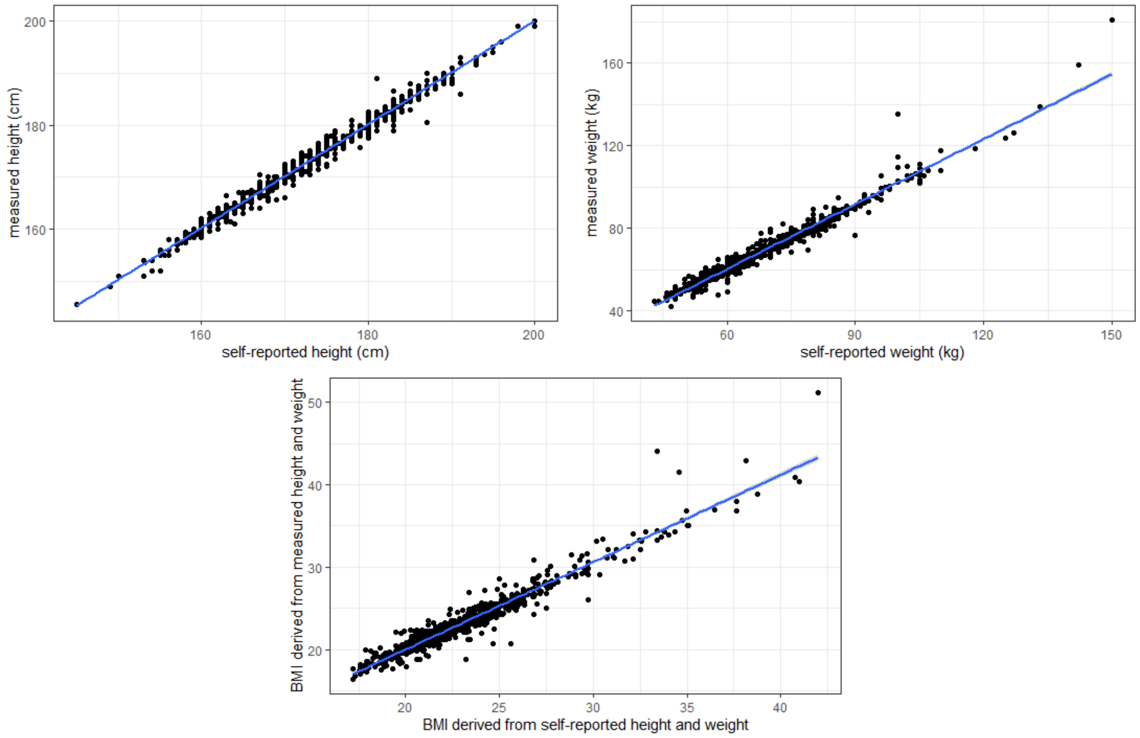


**Self-reported height, weight and BMI**

| | | height | | weight | | BMI | |
|---|---|---|---|---|---|---|---|
| | N | mean | sd | mean | sd | mean | sd |
| All | 752 | 171.5 | 9.5 | 68.6 | 14.2 | 23.2 | 3.6 |
| Male participants | 316 | 179.5 | 6.9 | 77.0 | 13.2 | 23.8 | 3.3 |
| Female participants | 436 | 165.7 | 6.3 | 62.5 | 11.6 | 22.7 | 3.8 |

**Measured height, weight and BMI**

| | | height | | weight | | BMI | |
|---|---|---|---|---|---|---|---|
| | N | mean | sd | mean | sd | mean | sd |
| All | 752 | 171.7 | 9.5 | 69.4 | 15.2 | 23.4 | 4.0 |
| Male participants | 316 | 179.6 | 7.1 | 77.6 | 14.8 | 24.0 | 3.9 |
| Female participants | 436 | 166.0 | 6.4 | 63.5 | 12.5 | 23.0 | 4.0 |

Pearson correlation coefficients were used to compare self-reported and measured height and weight. The body mass index (BMI) derived from the self-reported measurements (kg.m-2) was also compared with the BMI derived from the measured height and weight. Linear regressions were also used to model the relationship between self-reported (x) and measured (y) variables as (Δ).

$$y = \beta_0 + \beta_1 x + \varepsilon \qquad (\Delta)$$

Measured and self-reported height were highly correlated (r=0.99). Measured and self-reported weight also correlated strongly (r=0.98). The BMI derived from the self-reported measurements and the BMI derived from the clinical measurements were strongly correlated (r= 0.97). Linear regressions also showed high agreement between self-reported and measured height, weight, and BMI.

According to linear regressions, there seems to be a great match between self-reported height and measured height. Individuals with high weight tended to slightly underestimate their weight ($\beta_1$=1.048). The diagnostic of the residuals shows overall correct quality modeling for height and weight, but some individuals may have inflated the slope coefficient values. The outputs of the linear regressions are therefore to be considered with caution.

**Tables were generated using the Stargazer R package:**

Hlavac, Marek (2022). stargazer: Well-Formatted Regression and Summary Statistics Tables. R package version 5.2.3. https://CRAN.R-project.org/package=stargazer

| | *Dependent variable:* |
|---|---|
| | measured height |
| self-reported height ($\beta_1$) | $0.994^{***}$ |
| | (0.004) |
| Constant ($\beta_0$) | 1.167 |
| | (0.734) |
| Observations | 752 |
| $R^2$ | 0.986 |
| Adjusted $R^2$ | 0.986 |
| Residual Std. Error | 1.110 (df = 750) |
| F Statistic | $54{,}098.020^{***}$ (df = 1; 750) |
| *Note:* | $^{*}$p<0.1; $^{**}$p<0.05; $^{***}$p<0.01 |

| | *Dependent variable:* |
|---|---|
| | measured weight |
| self-reported weight ($\beta_1$) | $1.048^{***}$ |
| | (0.007) |
| Constant ($\beta_0$) | $-2.495^{***}$ |
| | (0.517) |
| Observations | 752 |
| $R^2$ | 0.964 |
| Adjusted $R^2$ | 0.964 |
| Residual Std. Error | 2.881 (df = 750) |
| F Statistic | $20{,}169.380^{***}$ (df = 1; 750) |
| *Note:* | $^{*}$p<0.1; $^{**}$p<0.05; $^{***}$p<0.01 |

| | *Dependent variable:* |
| --- | --- |
| | measured BMI |
| BMI from self-report ($\beta_1$) | 1.058*** |
| | (0.010) |
| Constant ($\beta_0$) | −1.138*** |
| | (0.237) |
| Observations | 752 |
| $R^2$ | 0.936 |
| Adjusted $R^2$ | 0.936 |
| Residual Std. Error | 1.009 (df = 750) |
| F Statistic | 11,037.050*** (df = 1; 750) |
| *Note:* | *p<0.1; **p<0.05; ***p<0.01 |

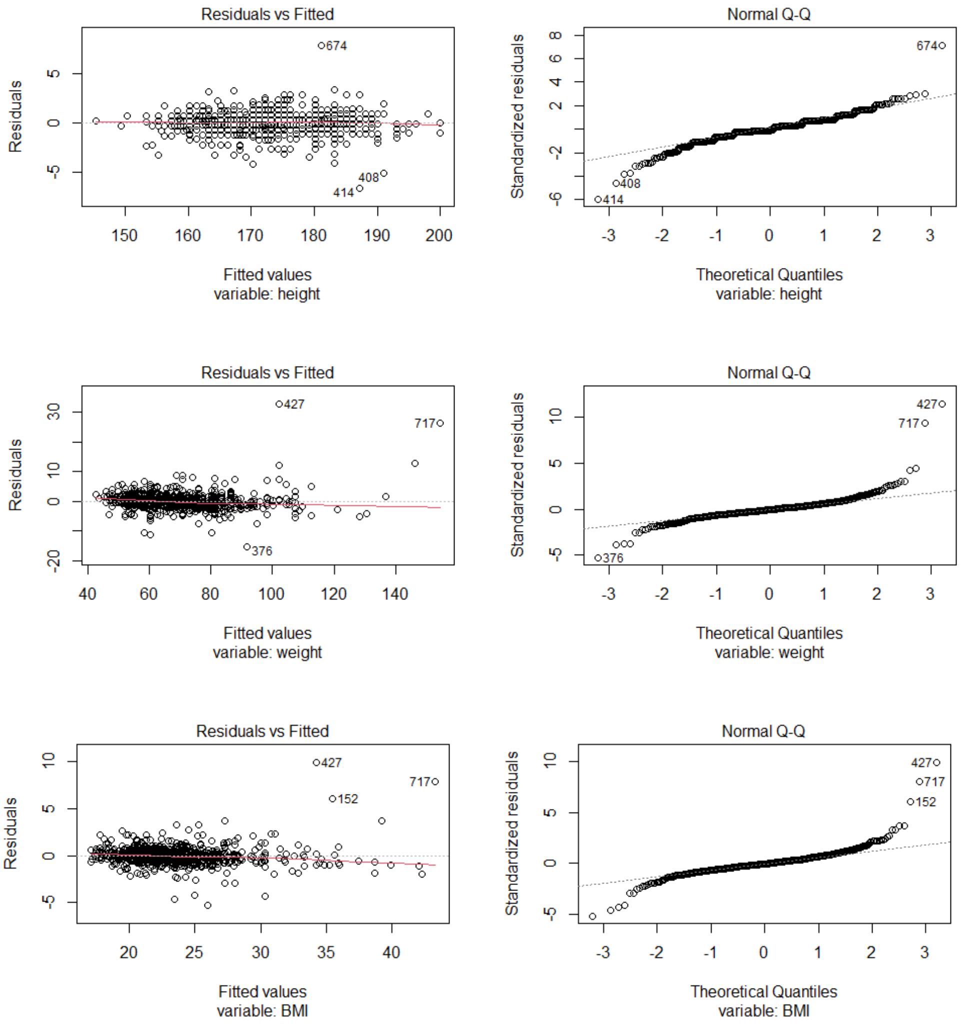
