## Additional file 4 for "Longitudinal multi-omics study reveals common etiology underlying association between plasma proteome and BMI trajectories in adolescent and young adult twins"

**Supplementary document**

### **1- Genotyping and polygenic score calculation in NTR**

###### Genotyping and imputation

Genotyping in the NTR was done on multiple platforms over time, namely Perlegen-Affymetrix, Affymetrix 6.0, Affymetrix Axiom, Illumina Human Quad Bead 660, Illumina Omni 1M and Illumina GSA. On each platform, genotyping was performed following manufacturers protocols, using the then appropriate calling software. As the SNPs of the Perlegen-Affymetrix, Illumina Human Quad Bead 660 and Illumina Omni 1M arrays were originally typed on older genome builds, these were lifted over to Build 37 HG19 based on RSid locations of the DBSNP 142 marker map. For each genotype platform, samples were removed if DNA sex did not match the expected phenotype, if the PLINK heterozygosity F statistic was < -0.10 or > 0.10, or if the genotyping call rate was < 0.90. SNPs were removed if the MAF < 1×10-6, if the Hardy-Weinberg equilibrium p-value was < 1×10-6, and/or if the call rate was < 0.95 (Purcell et al, 2007). Subsequently, for each platform, the genotype data was aligned with the 1000 Genomes reference panel using the HRC and 1000 Genomes checking tool (v4.3, <https://www.well.ox.ac.uk/~wrayner/tools/>), which tests and filters for SNPs with allele frequency differences larger than 0.20 as compared to the CEU population, palindromic SNPs and DNA strand issues (1000 Genomes Project Consortium et al, 2015).

The data of the six platforms was then merged into a single dataset, keeping all quality-controlled SNPs of each platform. For each individual, one platform was chosen in the following order: Affymetrix 6.0 > Illumina GSA > Affymetrix Axiom > Illumina Omni 1M > Illumina Human Quad Bead 660 > Perlegen-Affymetrix. Based on the ~10.8k SNPs that all platforms have in common, DNA Identity-By-Descent (IBD) state was estimated for all individual pairs using the PLINK (v1.9) and KING programs (Purcell et al, 2007; Manichaikul et al, 2010). These estimates were then compared to the expected familial relations, and samples were removed if these failed to fit. Post IBD check, a similar approach was used for DNA zygosity mismatches as well as samples having different DNA between platforms. CEU population outliers, based on per platform 1000 Genomes PC projection with the SMARTPCA software (v7), were removed from the data (Price et al, 2006).

Per platform, the data was phased using Eagle (v2.4.1, Loh et al, 2016; Delaneau et al, 2011) and then imputed to 1000 Genomes using Minimac (v2.0.1) following the Michigan imputation server protocols (Das et al, 2016). Post imputation, the resulting separate platform Variant Call Format (VCF) files were merged with BCFTOOLS into a single VCF file per chromosome, only for those SNPs present on all six platforms (Narasimhan et al, 2016). In the present study, genotyped participants were from the following platforms: Affymetrix 6.0 (N=573), Affymetrix Axiom (N=1), Illumina 660 Human beadchip (N=1), and Illumina GSA (N=1).

###### Computation of principal components

Twenty principal components (PCs) were re-calculated with SMARTPCA using LD-pruned 1000 Genomes–imputed SNPs that were also genotyped on at least one platform, had MAF > 0.05 and were not present in the long-range LD regions.

###### Polygenic score calculation

For the polygenic scoring, the imputed data were converted to best guess data and were filtered to include only ACGT SNPs, SNPs with MAF > 0.01, HWE p > 10 -5 and a genotype call rate > 0.98, and to exclude SNPs with more than 2 alleles. All mendelian errors were set to missing. The polygenic score (PGS) for BMI (Yengo et al, 2018) was calculated using summary statistics that omitted NTR from the discovery meta-analysis to avoid over-estimation of the genetic dispositions of BMI. Before calculating the PGS, linkage disequilibrium (LD) weighted betas were calculated from these summary statistics using the LDpred package to correct for the effects of LD and to maximize predictive accuracy of the PGS (Vilhjálmsson et al, 2015). The adjusted betas were calculated using a LD pruning window of 250 KB, and the infinitesimal prior (LDpred-inf). The 1000 genomes CEU population was used as reference sample used to estimate LD. The resulting betas were used to calculate the PGS using the PLINK (v1.90) software.
